# Prognostic role of COVID-19 pneumonia signs and other CT-biomarkers for survival in patients with malignant neoplasms: the ARILUS project

**DOI:** 10.1101/2025.11.14.25338295

**Authors:** A.A. Dyachenko, M.A. Bogdanov, D.V. Bogdanov, A.M. Grjibovski, A.A. Markov, E.A. Nazarova, A.A. Meldo, V.Yu. Chernina, M.G. Belyaev, V.A. Gombolevskiy, M.Yu. Valkov

## Abstract

**Background:** Clinically manifested pneumonia associated with COVID-19 infection in cancer patients has been associated with worse prognosis. The prognostic significance of subclinical manifestations of pulmonary infiltration is poorly understood.

**Aims:** To estimate the survival of cancer patients with signs of asymptomatic pneumonia detected by a multitarget artificial intelligence (AI) algorithm on chest computed tomography (CT) during the COVID-19 pandemic, and to evaluate factors associated with the risk of death in this population.

**Materials and methods:** This population-based cohort study, conducted within the ARILUS project, included 1,147 examinations of cancer patients performed between August 2020 and May 2021. Signs of pneumonia on CT were detected by the AI algorithm in 556 (48.4%) patients with malignant neoplasms (MN) who had no clinical manifestations of infection at the time of the examination. Overall survival (OS) was assessed using the life table and Kaplan-Meier methods. Multivariate Cox proportional hazards regression analysis was used to identify independent predictors of death and to assess the influence of potential confounding factors.

**Results:** During the follow-up period, 680 deaths were recorded among cancer patients, of which 575 deaths were due to MN progression. Three-year OS differed significantly between the groups: in patients without signs of pulmonary infiltration on CT, it was 60.7% (95% CI 56.6%– 64.7%), whereas in patients with AI-detected signs of pneumonia, it was 45.6% (95% CI 41.3%– 49.8%) (p<0.001). In the multivariable regression analysis, independent factors associated with an unfavorable prognosis (death) were: presence of pulmonary infiltration (adjusted hazard ratio (HR) 1.31; 95% CI 1.09–1.58), male sex (HR 1.25; 95% CI 1.00–1.57), and MN stage (HR 1.78 for stage II, HR 2.93 for stage III, and HR 4.74 for stage IV compared to stage I). Signs of pulmonary emphysema (HR 1.84), aortic aneurysm (HR 1.75), and coronary artery calcification (HR 1.22-1.34), which were significantly associated with the risk of death in the univariable analysis, lost their prognostic significance in the multivariable model after adjustment for other variables.

**Conclusions:** Asymptomatic COVID-19 pneumonia detected by AI on CT is an independent predictor of reduced overall survival in MN patients. Automated AI screening for such changes may be recommended in routine practice.

## Rationale

The emergence of the novel severe acute respiratory syndrome coronavirus 2 (SARS-CoV-2) triggered a global healthcare crisis, leading to shifts in standard approaches to the diagnosis and treatment of oncological diseases [1]. Cancer patients and healthcare professionals frequently encountered obstacles, as the benefits of specialized treatment had to be weighed against the increased risk of SARS-CoV-2 infection during hospitalization, higher frequency of outpatient visits, and the potential for more severe COVID-19 due to immunosuppression caused by antitumor therapy.

The mortality rate among cancer patients with symptomatic COVID-19 was 24–28% [2, 3], with the leading causes of death being respiratory distress syndrome and viral-associated pneumonia [4]. The extent of lung involvement on computed tomography (CT) serves as a predictor of worsening clinical progression of the viral infection [5, 6]. Moreover, the risk of pneumonia itself is higher in patients with malignant diseases [7], and the presence of comorbid cardiovascular conditions in cancer patients exacerbates the course of infection [8]. COVID-19-associated pneumonia may remain asymptomatic in a significant proportion of patients, with up to 50% of contacts with symptomatic pneumonia patients testing positive [9, 10]. Meanwhile, the primary and most sensitive diagnostic method for COVID-19-associated pneumonia is CT [11], the diagnostic accuracy of which improves significantly with the use of artificial intelligence (AI) algorithms [12].

The ARILUS study [13] aims to identify predictors of cardiovascular and respiratory diseases, as well as signs of osteoporosis, in cancer patients using chest CT scans analyzed by a leading Russian AI algorithm [14], with the goal of correcting these factors to prevent non-oncological mortality. In our study, conducted under the ARILUS protocol during the pandemic, radiological signs of pneumonia (predominantly CT-1) were detected in half of the patients with malignant neoplasms. Among patients with lung, head and neck, and upper gastrointestinal cancers, the frequency of pneumonia detection on CT was 57–70%. The relative risk of detecting pneumonia was significantly increased by 25% in patients with signs of emphysema and by 24–61% in those with varying degrees of coronary calcification [1]. The prognostic significance of asymptomatic lung involvement, detectable only on CT, remains unknown.

Survival analysis of cancer patients using population-based data allows for an assessment of the organization of oncological care and tracking of outcomes in cancer patients with timely access to mortality records. The Population-Based Cancer Registry of the Arkhangelsk Region and the Nenets Autonomous Okrug (PBCR AO and NAO) has been maintaining cancer records for the past 25 years, with data characterized by high completeness and accuracy, enabling high-quality epidemiological research.

### Study Objective

To evaluate the survival of cancer patients with signs of COVID-19-associated pneumonia detected by a multitarget AI algorithm on chest CT scans and to assess factors influencing the risk of death in these patients.

## Methods

### Study Design

The present study is part of the ARILUS project - the **Ar**khangelsk Research on the Impact of Multitarget Artificial **I**ntelligence for Computed Tomography in Reducing Non-Oncological **L**ethal O**u**tcome**s** in Patients with Malignant Neoplasms (ARILUS). Case selection for analysis was described previously [15]. The initial pool consisted of 11,173 chest CT scans performed at the Arkhangelsk Regional Clinical Oncology Center from April 1, 2020 to December 31, 2021 during the COVID-19 pandemic.

The study included asymptomatic patients with negative SARS-CoV-2 PCR tests (nasopharyngeal swabs). CT scans were performed for two groups of indications: (1) routine oncological imaging for diagnosis clarification and staging of malignant processes, and (2) emergency examination to exclude viral pneumonia in hospitalized patients with clinical signs of COVID-19.

Exclusion criteria included CT scans with significant atelectasis, contrast-enhanced series, images with severe motion artifacts, reconstructions with slice thickness >1.5 mm, incomplete thoracic coverage, and cases with missing or incorrect individual insurance account numbers (SNILS) required for linkage with the cancer registry database. From the initial 7,639 chest CT examinations, 1,147 cases (15.0%) from August 2020 to May 2021 were included in the final analysis. Sequential exclusions were made for the following reasons: significant atelectasis - 124 examinations, intravenous contrast use - 2,106, major motion artifacts - 586, reconstruction thickness >1.5 mm - 3,268, incomplete scan coverage - 14, missing/incorrect SNILS - 394.

Thus, all cases in the final dataset contained both chest CT data and corresponding records in the PBCR AO and NAO, including patient identification codes, SNILS, age at CT examination, sex, residential settlement type, and ICD-10 cancer diagnosis codes.

### Algorithm for COVID-19-Associated Pneumonia Detection

For detection and quantification of pulmonary infiltrative changes (both qualitative and quantitative, including percentage involvement per lung) characteristic of COVID-19-associated viral pneumonia (ICD-10 code U07.1), we used AI-based software “Intelligent Radiology Assistants” (developed by IRA Labs LLC, Moscow, Russia). This medical device complies with technical specifications TU 58.29.32-001-44270315-2021 and holds Roszdravnadzor registration certificate № RZN 2024/22895.

During the pandemic, this algorithm was implemented in primary care facilities in Moscow for automated analysis of chest CTs, standardizing diagnostics and objectively assessing lung involvement [16]. Each AI output contained two CT series: DICOM images and structured reports in DICOM SR format. Preliminary validation using independent datasets from the Moscow Experiment [17] demonstrated algorithm accuracy of 0.98, sensitivity 0.94, and specificity 0.94.

This multitarget algorithm enables simultaneous detection of multiple thoracic pathologies on CT, including pulmonary emphysema, aortic and pulmonary trunk diameter measurements, coronary artery calcification, epicardial/paracardial fat volume, and bone mineral density of thoracic vertebrae. Appendix 1 provides characteristics of all evaluated biomarkers and results from independent external validation during the Innovative Technologies Experiment.

### Survival Analysis

In the final cohort, AI-detected pneumonia signs were present in 556 patients (48.4%). We compared overall survival (OS) and cancer-specific survival (CSS) between patients with and without pneumonia signs.

For OS analysis, the endpoint was all-cause mortality. For CSS analysis, the endpoint was death from cancer progression or treatment complications, with non-cancer deaths censored at event date. Living patients were censored at March 20, 2025 - two months prior to the last registry update. The PBCR AO and NAO database undergoes monthly updates through linkage with regional mortality records.

### Ethical Approval

The study was approved by the Local Ethics Committee of Northern State Medical University (Protocol №07/10-238, 2023).

### Statistical Analysis

Categorical variables were compared using Pearson’s χ^2^ test. Survival analysis employed life tables and Wilcoxon-Gehan tests for 3-year survival comparisons. Univariate survival functions were assessed using Mantel-Cox log-rank tests. Given baseline characteristic differences between pneumonia-positive and negative groups, we performed multivariate Cox proportional hazards regression [18] to adjust for potential confounders. Statistical significance threshold was set at p<0.05. All survival estimates and hazard effects are reported with 95% confidence intervals (95% CI). Statistical analysis was performed using Stata v.18 (Stata Corp., TX, USA).

## Results

The distribution of patients by baseline characteristics, including clinical and demographic features along with AI analysis results from CT scans, is presented in Table 1.

**Table 1.** Characteristics of the groups.

| Feature | Total (n = 1147) | No evidence of COVID-19 pneumonia (n = 592) | COVID-19 pneumonia (n = 556) | p-value |
| --- | --- | --- | --- | --- |
| Sex |  |  |  |  |
| Female | 634 | 374 (59.0) | 260 (41.0) | <0.001 |
| Male | 513 | 218 (42.4) | 296 (57.6) |  |
| Age, years |  |  |  |  |
| 0-39 | 43 | 27 (62.8) | 16 (37.2) | 0.002 |
| 40-49 | 115 | 72 (62.6) | 43 (37.4) |  |
| 50-59 | 236 | 126 (53.4) | 110 (46.6) |  |
| 60-69 | 456 | 238 (52.2) | 218 (47.8) |  |
| 70-79 | 249 | 112 (45.0) | 137 (55.0) |  |
| 80 and older | 48 | 16 (33.3) | 32 (66.7) |  |
| **Place of residence** |  |  |  |  |
| Urban | 859 | 456 (53.1) | 403 (46.9) | 0.066 |
| Rural | 288 | 135 (46.9) | 153 (53.1) |  |
| **ICD-10 codes, cancers** |  |  |  |  |
| C00-14, C30-32, Head and neck cancers | 46 | 18 (39.1) | 28 (60.9) | <0.001 |
| C15, C16, Upper GIT cancers | 140 | 60 (42.9) | 80 (57.1) |  |
| C18-25, Lower GIT cancers | 238 | 148 (62.2) | 90 (37.8) |  |
| C34, Lung cancer | 185 | 55 (29.7) | 130 (70.3) |  |
| C43-49, Skin and soft tissues cancer | 74 | 37 (50.0) | 37 (50.0) |  |
| C50, Breast cancer | 151 | 79 (52.3) | 72 (47.7) |  |
| C51-58, Female genital organs cancers | 122 | 80 (65.6) | 42 (34.4) |  |
| C61, Prostate cancer | 50 | 29 (58.0) | 21 (42.0) |  |
| C64-68, Urinary system cancers | 76 | 39 (51.3) | 37 (48.7) |  |
| Other neoplasms | 65 | 46 (70.8) | 19 (29.2) |  |
| **Lung emphysema** |  |  |  |  |
| No | 1063 | 573 (53.9) | 490 (46.1) | <0.001 |
| Yes | 84 | 18 (21.4) | 66 (78.6) |  |
| **Aortic aneurysm** |  |  |  |  |
| No | 1129 | 583 (51.6) | 546 (48.4) | 0.545 |
| Yes | 18 | 8 (44.4) | 10 (55.6) |  |
| **Epicardial fat volume** |  |  |  |  |
| <125 ml | 519 | 238 (45.9) | 281 (54.1) | 0.254 |
| ≥125 ml | 370 | 184 (49.7) | 186 (50.3) |  |
| **Pulmonary trunk dilation** |  |  |  |  |
| No | 821 | 446 (54.3) | 375 (45.7) | 0.003 |
| Yes | 326 | 145 (44.5) | 181 (55.5) |  |
| **Coronary artery calcification, Agatston index** |  |  |  |  |
| 0 | 569 | 355 (62.4) | 214 (37.6) | <0.001 |
| 1-99 | 264 | 133 (50.4) | 131 (49.6) |  |
| 100-299 | 139 | 48 (34.5) | 91 (65.5) |  |
| ≥300 | 175 | 55 (31.4) | 120 (68.6) |  |
| **Osteoporosis, osteopenia** |  |  |  |  |
| No | 215 | 121 (20.9) | 94 (17.2) | 0.12 |
| Osteopenia | 437 | 230 (39.7) | 207 (37.9) |  |
| Osteoporosis | 473 | 228 (39.4) | 245 (44.9) |  |

**Notes:** CT - computed tomography, COVID-19 - COroNaVirus Disease 2019, ICD-10 - international classification of diseases 10th revision, GIT - gastrointestinal tract. Pathological signs of aneurysm and aortic dilatation, pulmonary trunk dilation, coronary calcification, and osteopenia were not determined in all CT series, as either the corresponding organs were not fully represented in these series or were not detected due to algorithm-specific characteristics.

Demographic characteristics and AI-identified CT findings were unevenly distributed among the compared subgroups of oncological patients. The likelihood of detecting CT findings pathognomonic for COVID-19-associated viral pneumonia was significantly higher in male patients, elderly individuals, those with head and neck malignancies, esophageal and gastric cancers, lung cancer, as well as in cases revealing CT evidence of pulmonary emphysema, pulmonary trunk dilation, significant coronary artery calcification (Agatston index ≥100), osteopenia and osteoporosis.

During the observation period following study inclusion, 680 deaths were recorded among cancer patients, including 575 deaths from malignant neoplasms, 42 from cardiovascular diseases, 44 from respiratory diseases, and 19 from other causes. The 1- and 3-year overall survival (OS) rates were 70.2% (95% CI 67.5%-72.7%) and 48.3% (95% CI 45.4%-51.1%) respectively. The 1- and 3-year cancer-specific survival (CSS) rates were 74.2% (95% CI 71.5%- 76.6%) and 49.0% (95% CI 46.0%-51.9%) respectively.

The presence of pneumonia signs associated with viral infection was linked to worse outcomes in both OS and CSS. The 1- and 3-year OS rates in patients without pneumonia signs were 80.8% (95% CI 77.4%-83.8%) and 60.7% (95% CI 56.6%-64.7%) respectively, compared to 65.9% (95% CI 61.7%-69.7%) and 45.6% (95% CI 41.3%-49.8%) in those with pneumonia signs (p<0.001). The detection of pulmonary infiltration was also associated with worse CSS outcomes: 65.9% (95% CI 61.7%-69.7%) and 45.6% (95% CI 41.3%-49.8%) survived beyond 1 and 3 years respectively, compared to 80.8% (95% CI 77.4%-83.8%) and 60.7% (95% CI 56.6%-64.7%) in patients without lung involvement (Figure 1).

**Figure 1.**
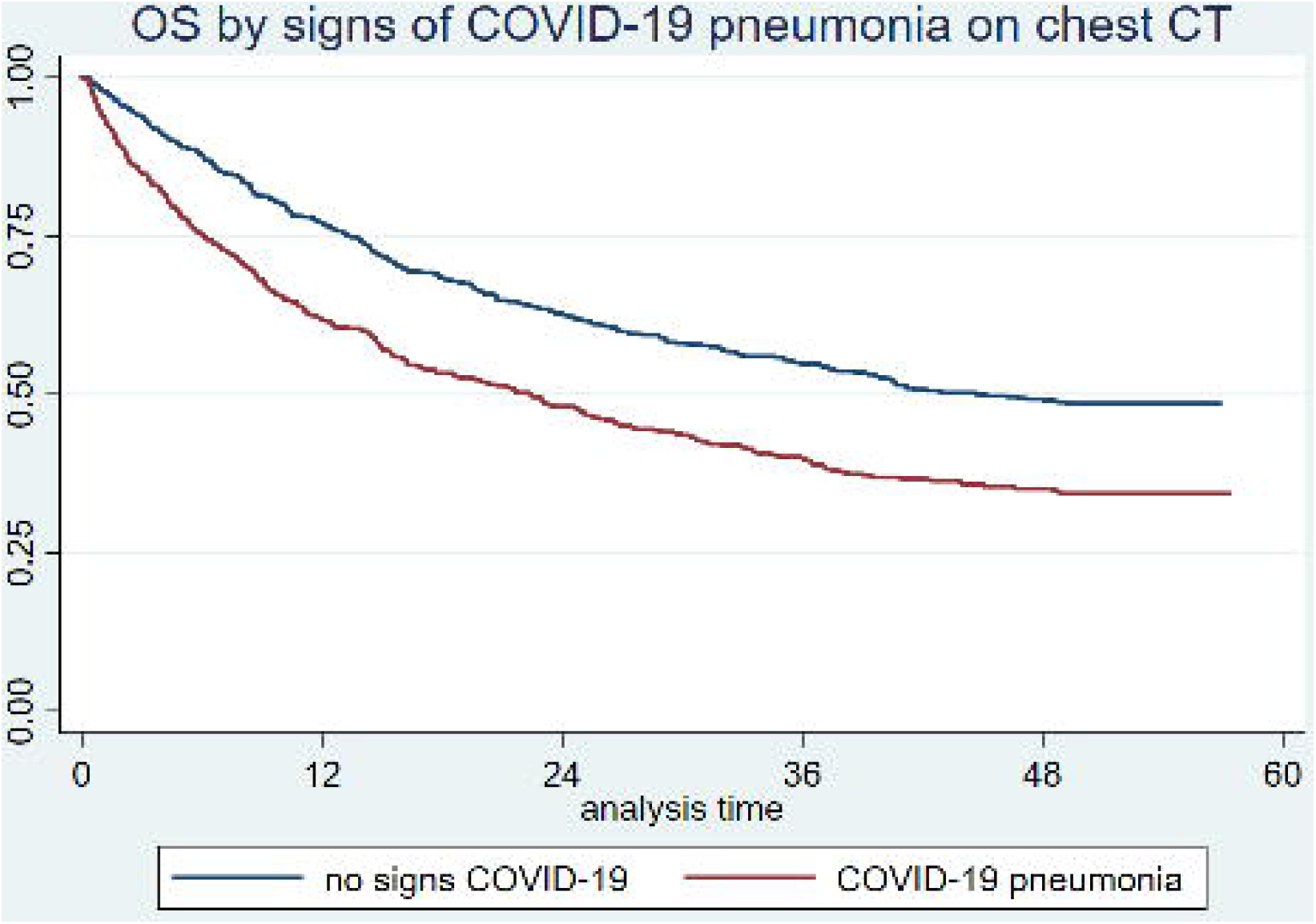
Overall (a) and cancer-specific (b) survival rates depending on the presence or absence of signs of COVID-19-associated pneumonia among cancer patients undergoing chest CT in 2020-2021 at the ACOD.

In general, the presence of COVID-19-associated pneumonia signs adversely affected overall survival prognosis in the majority of analyzed subgroups. In cases where pulmonary infiltration was detected, 3-year OS was statistically significantly lower among patients of both sexes, across all age groups, in both urban and rural residents, as well as in upper gastrointestinal tract malignancies, lung cancer, and urinary tract tumors. A less pronounced, statistically insignificant survival reduction was observed in head and neck cancers, breast cancer, female reproductive system tumors, and prostate cancer. At the same time, no significant impact of this factor on survival outcomes was detected in malignancies of lower gastrointestinal tract, skin and soft tissues.

When pulmonary infiltration signs were identified on CT, 3-year OS rates were lower both in the presence and absence of emphysema, with epicardial fat volume below and above 125 ml, with and without pulmonary trunk dilation signs, with and without coronary artery calcification, and in patients with and without signs of osteopenia and osteoporosis in thoracic vertebrae. Cancer patients without aortic aneurysm showed 11.6% lower 3-year OS when pulmonary infiltration was present (p<0.001), while no decrease was observed in 10 patients with COVID-19 signs compared to 8 patients without pulmonary infiltration in the aneurysm subgroup.

The detection of COVID-19 signs on CT significantly worsened prognosis more in male patients compared to females (p=0.021) and in patients aged 80 years and older (p=0.0001). Three-year OS rates in patients with pulmonary infiltration signs were statistically significantly lower when emphysema signs were present on CT - 22.7% (95% CI 13.5%-33.4%) versus 42.5% (95% CI 38.0%-46.8%), p=0.005 when absent, as well as when coronary artery calcification signs were present - 36.8% (95% CI 31.8%-41.9%), compared to absence of calcification signs - 45.3% (95% CI 38.6%-51.8%), p=0.042.

The results of univariable and multivariable Cox regression analysis assessing all-cause mortality risks in cancer patients with and without CT-detected COVID-19 infection signs, as well as depending on other baseline demographic characteristics and AI-identified CT findings, are presented in Table 3.

**Table 2.**
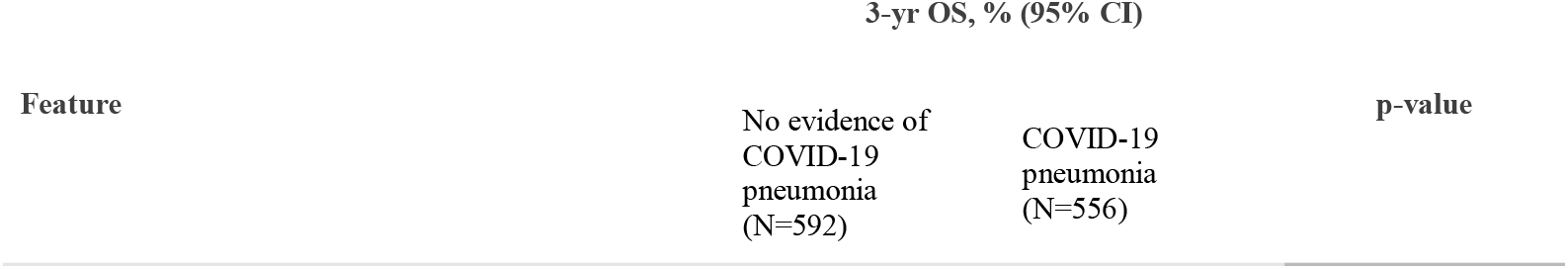
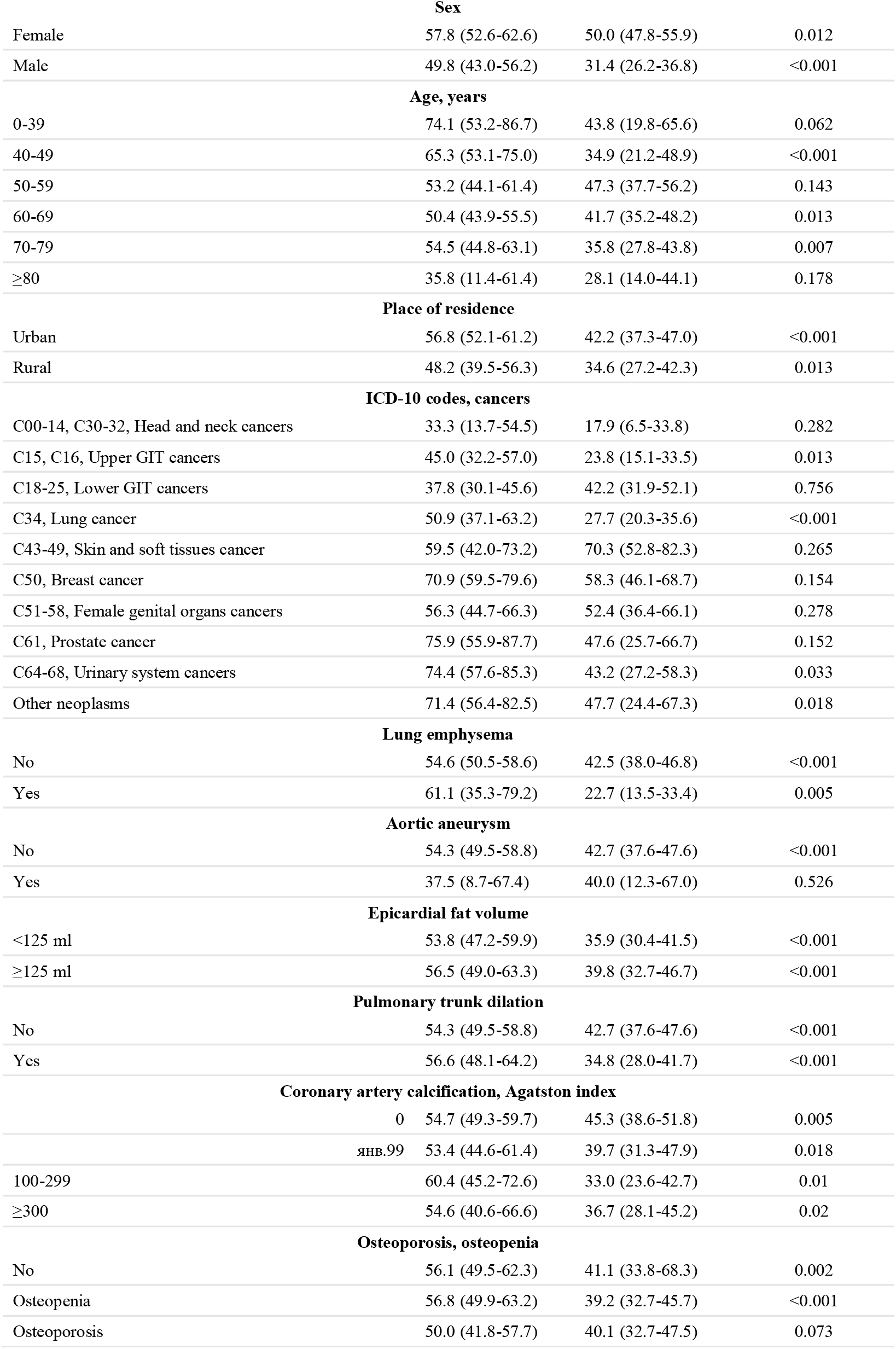
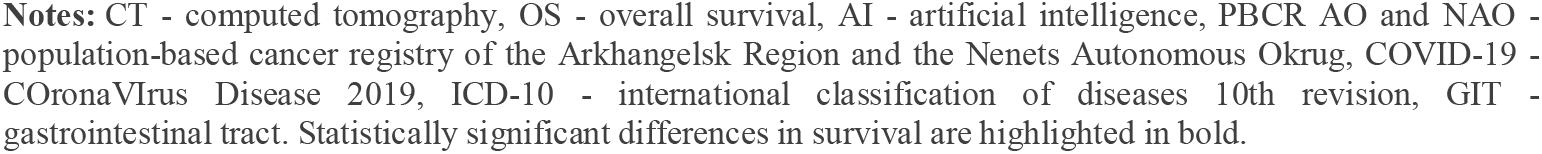
Overall survival rates for specific patient subgroups are presented in Table 2.

| Feature | 3-yr OS, % (95% CI) |  | p-value |
| --- | --- | --- | --- |
|  | No evidence of COVID-19 pneumonia (N=592) | COVID-19 pneumonia (N=556) |  |
|  | **Sex** |  |  |
| Female | 57.8 (52.6-62.6) | 50.0 (47.8-55.9) | 0.012 |
| Male | 49.8 (43.0-56.2) | 31.4 (26.2-36.8) | <0.001 |
|  | **Age, years** |  |  |
| 0-39 | 74.1 (53.2-86.7) | 43.8 (19.8-65.6) | 0.062 |
| 40-49 | 65.3 (53.1-75.0) | 34.9 (21.2-48.9) | <0.001 |
| 50-59 | 53.2 (44.1-61.4) | 47.3 (37.7-56.2) | 0.143 |
| 60-69 | 50.4 (43.9-55.5) | 41.7 (35.2-48.2) | 0.013 |
| 70-79 | 54.5 (44.8-63.1) | 35.8 (27.8-43.8) | 0.007 |
| ≥80 | 35.8 (11.4-61.4) | 28.1 (14.0-44.1) | 0.178 |
|  | **Place of residence** |  |  |
| Urban | 56.8 (52.1-61.2) | 42.2 (37.3-47.0) | <0.001 |
| Rural | 48.2 (39.5-56.3) | 34.6 (27.2-42.3) | 0.013 |
|  | **ICD-10 codes, cancers** |  |  |
| C00-14, C30-32, Head and neck cancers | 33.3 (13.7-54.5) | 17.9 (6.5-33.8) | 0.282 |
| C15, C16, Upper GIT cancers | 45.0 (32.2-57.0) | 23.8 (15.1-33.5) | 0.013 |
| C18-25, Lower GIT cancers | 37.8 (30.1-45.6) | 42.2 (31.9-52.1) | 0.756 |
| C34, Lung cancer | 50.9 (37.1-63.2) | 27.7 (20.3-35.6) | <0.001 |
| C43-49, Skin and soft tissues cancer | 59.5 (42.0-73.2) | 70.3 (52.8-82.3) | 0.265 |
| C50, Breast cancer | 70.9 (59.5-79.6) | 58.3 (46.1-68.7) | 0.154 |
| C51-58, Female genital organs cancers | 56.3 (44.7-66.3) | 52.4 (36.4-66.1) | 0.278 |
| C61, Prostate cancer | 75.9 (55.9-87.7) | 47.6 (25.7-66.7) | 0.152 |
| C64-68, Urinary system cancers | 74.4 (57.6-85.3) | 43.2 (27.2-58.3) | 0.033 |
| Other neoplasms | 71.4 (56.4-82.5) | 47.7 (24.4-67.3) | 0.018 |
|  | **Lung emphysema** |  |  |
| No | 54.6 (50.5-58.6) | 42.5 (38.0-46.8) | <0.001 |
| Yes | 61.1 (35.3-79.2) | 22.7 (13.5-33.4) | 0.005 |
|  | **Aortic aneurysm** |  |  |
| No | 54.3 (49.5-58.8) | 42.7 (37.6-47.6) | <0.001 |
| Yes | 37.5 (8.7-67.4) | 40.0 (12.3-67.0) | 0.526 |
|  | **Epicardial fat volume** |  |  |
| <125 ml | 53.8 (47.2-59.9) | 35.9 (30.4-41.5) | <0.001 |
| ≥125 ml | 56.5 (49.0-63.3) | 39.8 (32.7-46.7) | <0.001 |
|  | **Pulmonary trunk dilation** |  |  |
| No | 54.3 (49.5-58.8) | 42.7 (37.6-47.6) | <0.001 |
| Yes | 56.6 (48.1-64.2) | 34.8 (28.0-41.7) | <0.001 |
|  | **Coronary artery calcification, Agatston index** |  |  |
| 0 | 54.7 (49.3-59.7) | 45.3 (38.6-51.8) | 0.005 |
| январь.99 | 53.4 (44.6-61.4) | 39.7 (31.3-47.9) | 0.018 |
| 100-299 | 60.4 (45.2-72.6) | 33.0 (23.6-42.7) | 0.01 |
| ≥300 | 54.6 (40.6-66.6) | 36.7 (28.1-45.2) | 0.02 |
|  | **Osteoporosis, osteopenia** |  |  |
| No | 56.1 (49.5-62.3) | 41.1 (33.8-48.3) | 0.002 |
| Osteopenia | 56.8 (49.9-63.2) | 39.2 (32.7-45.7) | <0.001 |
| Osteoporosis | 50.0 (41.8-57.7) | 40.1 (32.7-47.5) | 0.073 |

**Notes:** CT - computed tomography, OS - overall survival, AI - artificial intelligence, PBCR AO and NAO - population-based cancer registry of the Arkhangelsk Region and the Nenets Autonomous Okrug, COVID-19 - COronaVirus Disease 2019, ICD-10 - international classification of diseases 10th revision, GIT - gastrointestinal tract. Statistically significant differences in survival are highlighted in bold.

**Table 3.** An univariable and multivariable Cox regression analysis of the OS survival depending on the initial clinical and demographic characteristics and findings of AI on the chest CT over the period 2020-2021.

| Feature | Univariable |  | Multivariable |  |
| --- | --- | --- | --- | --- |
|  | HR (95% CI) | p-value | HR (95% CI) | p-value |
| **COVID-19 signs on chest CT** |  |  |  |  |
| No | 1.00 (ref.) |  | 1.00 (ref.) |  |
| Yes | 1.54 (1.32-1.80) | <0.001 | 1.31 (1.09-1.58) | 0.004 |
| **Sex** |  |  |  |  |
| Female | 1.00 (ref.) |  | 1.00 (ref.) |  |
| Male | 1.63 (1.40-1.89) | <0.001 | 1.25 (1.00-1.57) | 0.048 |
| **Age, years** |  |  |  |  |
| 0-39 | 1.00 (ref.) |  | 1.00 (ref.) |  |
| 40-49 | 1.17 (0.70-1.98) | 0.538 | 0.85 (0.48-1.48) | 0.558 |
| 50-59 | 1.37 (0.85-2.23) | 0.197 | 0.84 (0.49-1.44) | 0.527 |
| 60-69 | 1.57 (0.99-2.50) | 0.058 | 0.92 (0.53-1.57) | 0.747 |
| 70-79 | 1.67 (1.04-2.69) | 0.035 | 1.00 (0.57-1.76) | 0.991 |
| ≥80 | 2.13 (1.22-3.72) | 0.008 | 1.29 (0.66-2.51) | 0.456 |
| **Place of residence** |  |  |  |  |
| Urban | 1.00 (ref.) |  | 1.00 (ref.) |  |
| Rural | 1.34 (1.14-1.59) | 0.001 | 1.19 (0.98-1.44) | 0.082 |
| **Stage\*** |  |  |  |  |
| I | 1.00 (ref.) |  | 1.00 (ref.) |  |
| II | 1.44 (1.06-1.95) | 0.018 | 1.78 (1.25-2.54) | 0.01 |
| III | 2.54 (1.94-3.32) | <0.001 | 2.93 (2.14-4.02) | <0.001 |
| IV | 4.38 (3.37-5.70) | <0.001 | 4.74 (3.49-6.45) | <0.001 |
| Not known | 1.85 (0.89-3.84) | 0.1 | 3.20 (1.36-7.52) | 0.007 |
| **Lung emphysema** |  |  |  |  |
| No | 1.00 (ref.) |  | 1.00 (ref.) |  |
| Yes | 1.84 (1.42-2.38) | <0.001 | 1.29 (0.95-1.75) | 0.105 |
| **Aortic aneurysm** |  |  |  |  |
| No | 1.00 (ref.) |  | 1.00 (ref.) |  |
| Yes | 1.75 (1.05-2.92) | 0.032 | 1.20 (0.66-2.17) | 0.553 |
| **Epicardial fat volume** |  |  |  |  |
| <125 ml | 1.00 (ref.) |  | 1.00 (ref.) |  |
| ≥125 ml | 0.95 (0.80-1.13) | 0.563 | 0.94 (0.78-1.14) | 0.54 |
| **Coronary artery calcification, Agatston index** |  |  |  |  |
| 0 | 1.00 (ref.) |  | 1.00 (ref.) |  |
| 1-99 | 1.22 (1.02-1.49) | 0.034 | 0.83 (0.65-1.04) | 0.106 |
| 100-299 | 1.44 (1.14-1.81) | 0.002 | 0.99 (0.75-1.30) | 0.935 |
| ≥300 | 1.34 (1.08-1.66) | 0.009 | 0.81 (0.61-1.06) | 0.122 |
| **Osteoporosis, osteopenia** |  |  |  |  |
| No | 1.00 (ref.) |  | 1.00 (ref.) |  |
| Osteopenia | 1.12 (0.94-1.35) | 0.201 | 1.03 (0.84-1.28) | 0.742 |
| Osteoporosis | 1.25 (1.03-1.51) | 0.022 | 1.06 (0.84-1.34) | 0.626 |

**Notes:** HR - hazard ratio, CT - computed tomography, COVID-19 - COronaVirus Disease 2019, CI - confidence intervals, ref. - reference category. \*Although within each ICD-10 category stage was an independent predictor of death from any cause, no adjustment was made for uneven distribution of stage among topographies. Statistically significant differences in HR are highlighted in bold.

In the univariable analysis, the presence of pulmonary infiltration signs detected by AI-based CT processing was associated with a 54% increased risk of death from malignant neoplasms. After adjustment for all available factors, this risk remained significant with an HR of 1.31 (95% CI 1.09-1.58).

In the multivariable model incorporating all available variables through forced entry, additional independent predictors of poor prognosis included male sex (HR 1.25) and disease stage (HR 1.78, 2.93, and 4.74 for stages II, III, and IV respectively, compared to stage I). The initially elevated relative risk of death observed in patients over 70 years old became comparable to other age subgroups in the adjusted analysis. While rural residence showed a 19% increased mortality risk in preliminary analysis, this association lost statistical significance in the multivariate model.

Markers of potentially fatal non-oncological pathology identified by AI-based CT analysis demonstrated significant associations with mortality risk in univariable analysis: pulmonary emphysema increased risk by 84%, aortic aneurysm by 75%, coronary artery calcification by 22-44% (varying by severity), and osteoporosis by 25%. However, these associations became statistically insignificant after adjustment for other factors in the multivariable analysis.

## Discussion

### Summary of the main study findings

Our population-based study demonstrated that detection of pneumonia signs on chest CT scans of cancer patients examined and treated during the COVID-19 pandemic at the Arkhangelsk Regional Oncology Center was associated with a 31% increased mortality risk. This adverse prognostic effect was observed across all age and sex subgroups, regardless of residence (urban/rural). Pulmonary infiltration signs correlated with significantly reduced 3-year OS in upper GI tract, lung, and urinary tract malignancies. Notably, among all AI-identified CT biomarkers, this was the only pathology that maintained significant impact on overall survival in multivariate analysis. Initially significant predictors - emphysema, aortic aneurysm, coronary calcification, and osteoporosis signs - lost their prognostic value after full adjustment.

### Discussion of the main findings

Early pandemic data established that cancer patients faced higher SARS-CoV-2 infection risks versus general population, even without active treatment [19,20]. Those with COVID-19 symptoms showed greater severe outcome risks, particularly post-chemotherapy/surgery patients [21]. Key poor prognosis factors included age, male sex, and comorbidities [22]. Mortality risks in asymptomatic cancer patients remain understudied, primarily in short-term, small cohorts [23,24].

Our prior analysis revealed pulmonary involvement in 50% of asymptomatic, PCR-negative cancer patients - a hospitalization/prerequisite during the pandemic. Higher risks occurred in lung, head/neck, upper GI, and breast cancers, plus cases with concurrent emphysema/coronary calcification [15]. However, in current survival analysis, CT biomarkers lost prognostic significance after adjustment. Significantly reduced 3-year survival with COVID-19 signs in lung/upper GI cancers confirms both their susceptibility and vulnerability to viral infection, aligning with UKCCMP data showing 51% increased mortality with respiratory/intrathoracic tumors and severe COVID-19 [22].

Male sex emerged as an independent mortality predictor (25% increased risk). Symptomatic male cancer patients showed 63% higher 30-day mortality [25], while TERAVOLT identified smoking (OR 3.2) as sole multivariate risk factor [26] - more prevalent among males. Our data reveal elevated male mortality risk even without COVID-19 symptoms, potentially explained by higher smoking rates, comorbidities, testosterone deficiency, and estrogen’s immunoprotective role, though meta-analyses show conflicting evidence [27].

We used overall observed survival as our endpoint. While Russia’s death registration system (Federal Law #143-FZ, 15.11.1997) [28] is robust, cancer death cause accuracy may be compromised by insufficient death certificate detail, absent autopsies, and administrative factors. Misclassification rates range 15-48%, often erroneously attributing deaths to cancer [29,30]. CT-detected pneumonia increased all-cause mortality risk, potentially reflecting COVID-19’s disease course impact (including care delays) and viral immunosuppression.

The PBCR AO/NAO meets international standards, validated in global studies. In the Cancer Incidence in Five Continents (Vol. XII) [31] data quality indicators were at acceptable level: morphological verification (MV%) was 84.2% in men and 86.3% in women. The death-certificate-only cases (DCO%) were 8.5% (men), 7.3% (women). High MV% confirms registry reliability, while low DCO% reflects comprehensive case capture.

AI algorithms now enable automated chest CT interpretation in post-COVID cancer patients, detecting infiltrates and assessing prognostic value. UK Coronavirus Cancer Monitoring Project found persistent high mortality risk with moderate CT changes in asymptomatic/mild COVID-19 solid tumor patients, especially elderly males [32]. Chinese/US studies confirm automated lung severity scores independently predict poor outcomes even without symptoms [21], supporting AI integration for risk stratification and treatment adaptation.

### Study strengths

This study features a large representative cohort (n=1147) linked to a high-quality regional cancer registry, minimizing loss-to-follow-up and ensuring outcome validity. Standardized AI-CT analysis reduces interoperator variability and enhances reproducibility. Uniform data collection protocols establish a robust framework for implementing AI screening of subclinical COVID-19 pneumonia in routine oncology practice.

### Limitations

While retrospective and regionally confined, the population-based design and rigorous registry maintenance mitigate selection bias. Over 70% of initial CTs were excluded due to contrast/artifacts, but this enhanced image homogeneity and analysis reliability. Scanning parameters varied (slice thickness ≤1.5mm), though the certified AI algorithm was pretrained on heterogeneous data and demonstrates adequate sensitivity for subcentimeter changes.

Different COVID-19 variants may confer varying mortality risks. According to Arkhangelsk Regional Rospotrebnadzor, genotyping began in 2021 (Order #56, 19.02.2021), sequentially identifying 2021: Beta (South African), B.1.617.2 (Delta) (2021) and Delta, Omicron in 2022.

Evidence suggests reinfection after prior immunity may follow milder courses [33]. Lack of vaccination data in the registry prevents assessment of its protective effect. Future linkage via SNILS will enable evaluation of variant/vaccination impacts - a focus of planned analyses.

## Conclusion

While cancer patients’ COVID-19 vulnerability is established - particularly with recent treatment, thoracic tumors, male sex, or advanced age - subclinical lung involvement’s mortality risk in asymptomatic/PCR-negative cases remained unclear.

Our study proves AI-detected CT pneumonia signs independently increase mortality risk by 31% across all demographics in asymptomatic/PCR-negative cancer patients during the pandemic.

This biomarker was the sole significant CT predictor in multivariate analysis, driving marked 3-year OS reductions in lung, upper GI, and urinary tract cancers. Male sex independently increased mortality risk (25%) even without COVID-19 symptoms.

These population-level registry data first demonstrate AI-identified subclinical pulmonary infiltration as a significant vulnerability marker regardless of viral symptoms. Clinically, they justify integrating AI-CT analysis into routine oncology workups - especially for males and specified tumor sites-enabling risk stratification and timely management adjustments (intensified monitoring, preventive COVID-19 therapy, oncology treatment adaptation). Scientifically, they validate automated CT assessment as a reliable prognostic tool while highlighting needs for variant/vaccination impact studies - planned in subsequent analyses.

## Data Availability

All data produced in the present study are available upon reasonable request to the authors

https://www.arilus.ru

## Author Contributions

Dyachenko A.A. – research concept development, methodology design, work analysis, manuscript drafting, critical text revision, results interpretation, final manuscript approval.

Grzybowski A.M. – methodology design, statistical analysis and interpretation, critical text revision, results interpretation.

Markov A.A. – work analysis, critical text revision, results interpretation.

Bogdanov M.A. – work analysis, critical text revision, results interpretation.

Bogdanov D.V. – work analysis, critical text revision, results interpretation.

Chernina V.Yu. – work analysis, critical text revision, results interpretation.

Nazarova E.A. – work analysis, critical text revision, results interpretation.

Meldo A.A. – critical text revision, results interpretation.

Belyaev M.G. – critical text revision, results interpretation.

Gomboevsky V.A. – scientific supervision, research concept development, methodology development, critical text revision, results interpretation.

Valkov M.Yu. – scientific supervision, research concept development, methodology development, critical text revision, results interpretation.

All authors approved the final version of the article.

## Acknowledgments

The authors thank the staff of the Radiology Department at AKOD for their continuous and intensive CT data collection during the COVID-19 pandemic in the region. Special gratitude goes to the team of the Population-Based Cancer Registry for their over 20 years of unique work in systematizing data on cancer patients from two Russian regions.

